# Interventions to treat post-traumatic stress disorder (PTSD) in vulnerably housed populations and trauma-informed care: A scoping review

**DOI:** 10.1101/2021.06.07.21258494

**Authors:** Alexandria Bennett, Kien Crosse, Michael Ku, Nicole E. Edgar, Amanda Hodgson, Simon Hatcher

**Affiliations:** Clinical Epidemiology Program, Ottawa Hospital Research Institute, Ottawa, ON, Canada; Faculty of Medicine, University of Ottawa, Ottawa, ON, Canada; Health Sciences Library, University of Ottawa, Ottawa, ON, Canada; Department of Psychiatry, University of Ottawa, Ottawa, ON, Canada

**Author notes:** **Protocol registration**: Open Science Framework: https://osf.io/mpjgn. We acknowledge that all authors have contributed to this paper mandated by the International Committee of Medical Journal Editors. Corresponding author: Simon Hatcher, The Ottawa Hospital Riverside Campus, 1919 Riverside Drive, Suite 406, Ottawa, ON K1H 7W9.

**Keywords:** trauma-informed care, post-traumatic stress disorder, homelessness

## Abstract

**Objectives:** The goals of this study are to identify and analyse interventions that aim to treat Post Traumatic Stress Disorder (PTSD) and complex PTSD in individuals who are vulnerably housed and to describe how these treatments have been delivered using trauma-informed care.

**Design:** Scoping review

**Search strategy:** We searched electronic databases including MEDLINE, Embase, PsycINFO, CINAHL, the Cochrane Library, Web of Science, and PTSDpubs for published literature up to March 2020 for any studies that examined the treatment of PTSD in adults who were vulnerably housed. Websites of relevant organizations and other grey literature sources were searched to supplement the electronic database search. The characteristics and effect of the interventions were analyzed. We also explored how the interventions were delivered and the elements of trauma-informed care that were described.

**Results:** 26 studies were included. We identified four types of interventions: (1) trauma focused psychotherapies; (2) non-trauma psychotherapies; (3) housing interventions; and (4) pharmacotherapies. The trauma-informed interventions were small case series and the non-trauma focused therapies included four randomized controlled trials, were generally ineffective. Of the ten studies which described trauma-informed care the most commonly named elements were physical and emotional safety, the experience of feeling heard and understood, and flexibility of choice. The literature also commented on the difficulty of providing care to this population including lack of private space to deliver therapy; the co-occurrence of substance use; and barriers to follow-up including limited length of stay in different shelters and high staff turnover.

**Conclusions:** This scoping review identified a lack of high-quality trials to address PTSD in the vulnerably housed. There is a need to conduct well designed trials that take into account the unique setting of this population and which describe those elements of trauma-informed care that are most important and necessary.

**Strengths and limitations of this study:**

- First review of how trauma-informed care is being operationalized in the treatment of PTSD for the vulnerably housed.
- Provides a clear picture emphasizing the need to consistently utilize a trauma-informed care approach for PTSD treatment in the vulnerably housed.
- Largely homogenous populations (primarily women and US Veterans) included in studies, may not be representative of broader population.
- Often no mention of ethnic or cultural factors to consider when providing treatment.
- Many of the studies available were of low methodological quality.

## Background

Individuals who are vulnerably housed have higher mortality and morbidity rates compared to the general population.[1,2] A common vulnerability factor for many disorders is the experience of trauma. It is estimated that as many as 91% of individuals who are homeless have experienced at least one traumatic event[3] and up to 99% have experienced childhood trauma.[4,5] A recent qualitative study examining the pathways of men who become homeless long-term in Ontario, found that all of them had experienced complex childhood traumas.[6]

One consequence of exposure to trauma is either post-traumatic stress disorder (PTSD) or complex PTSD (cPTSD). PTSD results in re-experiencing the event, avoidance of reminders of the event, and persistent hypervigilance and awareness of threat. cPTSD results from prolonged threatening events which the individual cannot escape from (such as childhood abuse) and, in addition to PTSD symptoms, results in problems with affect regulation, negative beliefs about oneself, and difficulty in sustaining relationships.[7] Diagnosed PTSD rates in the homeless are significantly higher than the Canadian population, ranging between 21% and 53%[8–11] which may still be an underestimate of the actual prevalence.

Health care poses a unique and difficult challenge in the vulnerably housed, and a history of trauma, often perpetrated by people in caring roles such as parents or other family, makes accepting and engaging in treatment difficult.[6,12] Being vulnerably housed also exposes people to further traumas and re-victimization. [5,13,14] Further, PTSD is often poorly recognized as many vulnerably housed people with PTSD self-medicate with alcohol or other substances. There are also difficulties with accessing appropriate health care because of poverty and organizational barriers within health care providers.[15] These issues result in an underserved population with complex health needs that traditional mental health care is poorly equipped to serve.[10,16,17] Therefore, providing health care services to the vulnerably housed requires a degree of flexibility in terms of how services are provided, who provides them, when, and where. The approach recommended in clinical guidelines is trauma-informed care.[18–20]

The Substance Abuse and Mental Health Services Administration (SAMHSA) has defined trauma-informed care as a program, organization, or system that realizes the widespread impact of trauma and understands potential paths for healing; recognizes the signs and symptoms of trauma in staff, clients, and others involved with the system; and responds by fully integrating knowledge about trauma into policies, procedures, practices, and settings.[21] In 2010, Hopper described the four core principles of trauma-informed care in homeless services as trauma awareness, safety, choice and empowerment and a strengths based approach. The National Centre on Family Homelessness outlined several reasons why programs need to be trauma-informed,[22] including that trauma can impact how people access services, including viewing people and services as unsafe; recognition that people adapt to trauma to keep themselves safe including abusing substances, becoming aggressive, or withdrawing; and, programs and services cannot be “one size fits all”.

None of the five major Clinical Practice Guidelines for the treatment of PTSD[23] address the treatment of homeless people apart from the National Institute for Health and Care Excellence (NICE) guidance which states, “that methods of access to services take into account the needs of specific populations of people with PTSD, …including people who are homeless”. It doesn’t describe what these methods of access may be. A recent clinical practice guideline for the homeless and vulnerably housed does not include treatment for PTSD although it does recommend trauma-informed care without defining what this is.[20]

People who are vulnerably housed are unique amongst those who have PTSD because of the very high rates of exposure to trauma; the frequent use of substances to self-medicate symptoms; high rates of physical and mental comorbidities; the difficulty of forming caring relationships; and the difficulty of engaging with traditional health services often due to poverty and systemic barriers. We conducted this scoping review to explore the literature on what treatments have been used in this population and how trauma-informed care has been used to deliver these treatments.

### Objectives and rationale

This scoping review aims to provide an overview of the literature on the treatment of PTSD and cPTSD in individuals who are vulnerably housed, how these treatments have been delivered and, if trauma-informed care was used, how that was operationalized.

The specific research questions guiding this scoping review are:

1. What interventions are described in the literature for the treatment of PTSD and cPTSD in individuals who are vulnerably housed?
2. Are there any interventions for treatment of PTSD and cPTSD in the vulnerably housed which are described as trauma-informed, and how is this operationalized?

## Methods

We conducted a scoping review following the methodological framework proposed by Arksey and O’Malley [24] in addition to the methods manual published by the Joanna Briggs Institute’s Methodology for Scoping Reviews.[25] Our review also complies with the Preferred Reporting Items for Systematic Reviews and Meta-Analyses extension for Scoping Reviews (PRISMA-ScR) checklist.[26] The protocol is registered with the Open Science Framework (https://osf.io/mpjgn).[27]

### Information sources and literature search

A health librarian (AH) conducted electronic database searches in MEDLINE, Embase, PsycINFO (via Ovid), CINAHL (via EBSCO), The Cochrane Library (via Wiley), Web of Science and PTSDpubs (via Proquest) from inception until March 2020. The search was peer reviewed following the Peer Review of Electronic Search Strategies (PRESS) guidelines.[28] The full search strategy is available in an additional file (Appendix 1). No limits to language or publication date were applied. A thorough targeted search of the grey literature was conducted to identify any non-indexed studies including unpublished trial data, dissertations, theses, and conference proceedings. The Canadian Agency for Drugs and Technologies in Health (CADTH) Grey Matters Checklist was used to structure our grey literature search as well as identify key websites (i.e., organizations that focus on homelessness). We also hand-searched the reference list of identified reports for additional relevant studies that were not captured in the initial search.

### Study selection: inclusion criteria

We included published and unpublished primary research studies reporting any quantitative, qualitative, mixed-or multi-methods research which includes comparative and non-comparative methods evaluating an intervention that looks to treat PTSD in adults (18 years of age or older) who are vulnerably housed. For the purposes of this review, vulnerably housed populations are defined as those who are unsheltered, emergency sheltered, provisionally accommodated, and/or at risk of homelessness.[29] Study designs include randomized controlled trials (RCTs), cluster RCTs, quasi-experimental studies, cohort studies, cross-sectional/survey studies, case studies and controlled before and after studies. We excluded reviews, commentaries, and editorials.

### Study selection: Screening process

The citations from our comprehensive search strategy were uploaded into Covidence systematic review software (Veritas Health Innovation, Melbourne, Australia. Available at www.covidence.org). The screening process included phase I (titles and abstracts) and phase II (full texts) to identify relevant studies. In both phases, titles were screened in duplicate by two independent reviewers (AB, KC, MK) following the eligibility criteria previously outlined. Prior to each stage of screening, all reviewers ran pilot screening to identify and address any inconsistencies in applying the inclusion and exclusion criteria. Disagreements between reviewers were resolved by discussion and consultation of a third party (SH) if a consensus could not be reached.

### Data abstraction and charting

All included full-text studies were reviewed and abstracted by a single reviewer (AB) using a pilot-tested data abstraction form in Excel. Data that was abstracted included study characteristics, study design, population characteristics, details about the intervention and the trauma-informed approach, and any key findings. A secondary reviewer (KC, MK) verified the abstracted data and any discrepancies between reviewers were resolved through discussion.

The charting process included organizing and interpreting data by sifting, categorizing, and sorting material according to key issues and themes.[24] We charted the data based on whether the interventions to treat PTSD were individual or groups-based therapies and then again based on treatment type (i.e., cognitive-based therapy, pharmacological, contingency management, etc.).

We assessed evidence level by the approach outlined by Burns, Rohrich and Chung (2012) in The Levels of Evidence and their role in Evidence-Based Medicine (see Burns et al. 2012, p.10, Table 4 for Levels of Evidence for Therapeutic Studies).[30] We decided if the treatment was delivered by a trauma-informed approach by examining the components as outlined by Hooper et al (2010).[18] Studies were marked as “Yes” if they explicitly stated using a trauma-informed approach, while those that used components of a trauma-informed approach, but did not explicitly state a trauma-informed approach were marked as “partial.”

#### Patient Involvement

Persons with lived experience were included in the design stage of this project. Patients were not directly involved with the scoping review process. However, patients will be engaged for design and dissemination during the second phase of this project as informed by this scoping review.

## Results

The electronic search resulted in a total of 2,201 citations. We identified 1 study in our grey literature search. After de-duplication, 2,189 unique titles and abstracts were screened at phase I, of which 2,045 records were excluded, and 144 full-text articles were moved to phase II screening. A total of 26 articles met our inclusion criteria. The details of our selection process are illustrated in our PRISMA flow-chart in Figure 1.

**Figure 1.**
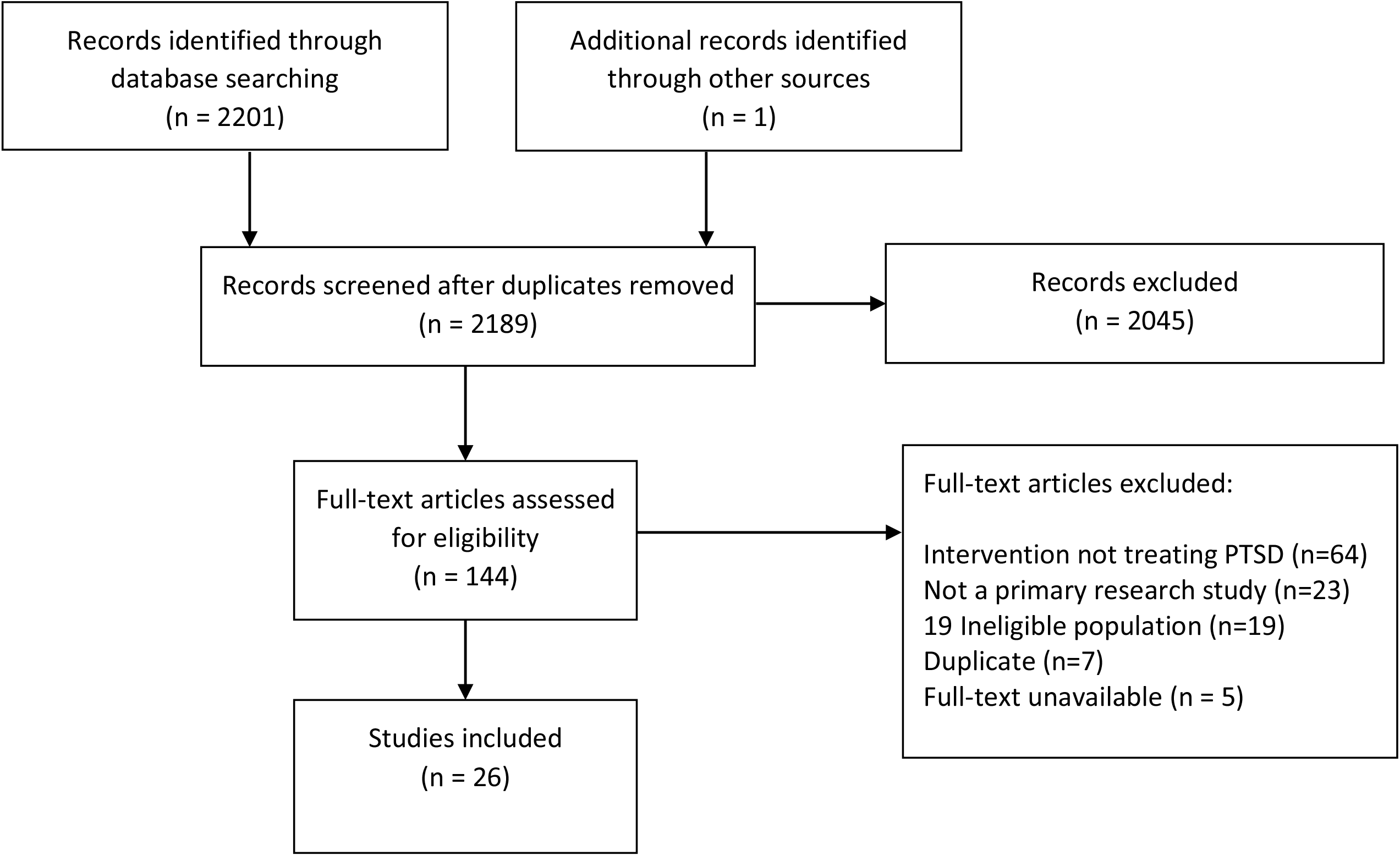
PRISMA flow-chart of included studies.

Most studies were conducted in the USA, apart from three studies which were from Canada, Spain, and the Netherlands published between the years 1999 and 2020. We identified 6 randomized-controlled trials (RCT), one quasi-RCT, 5 pre-post interventional studies, 4 case studies, and 3 pilot trials. A description of included studies is provided in Table 1.

**Table 1.**
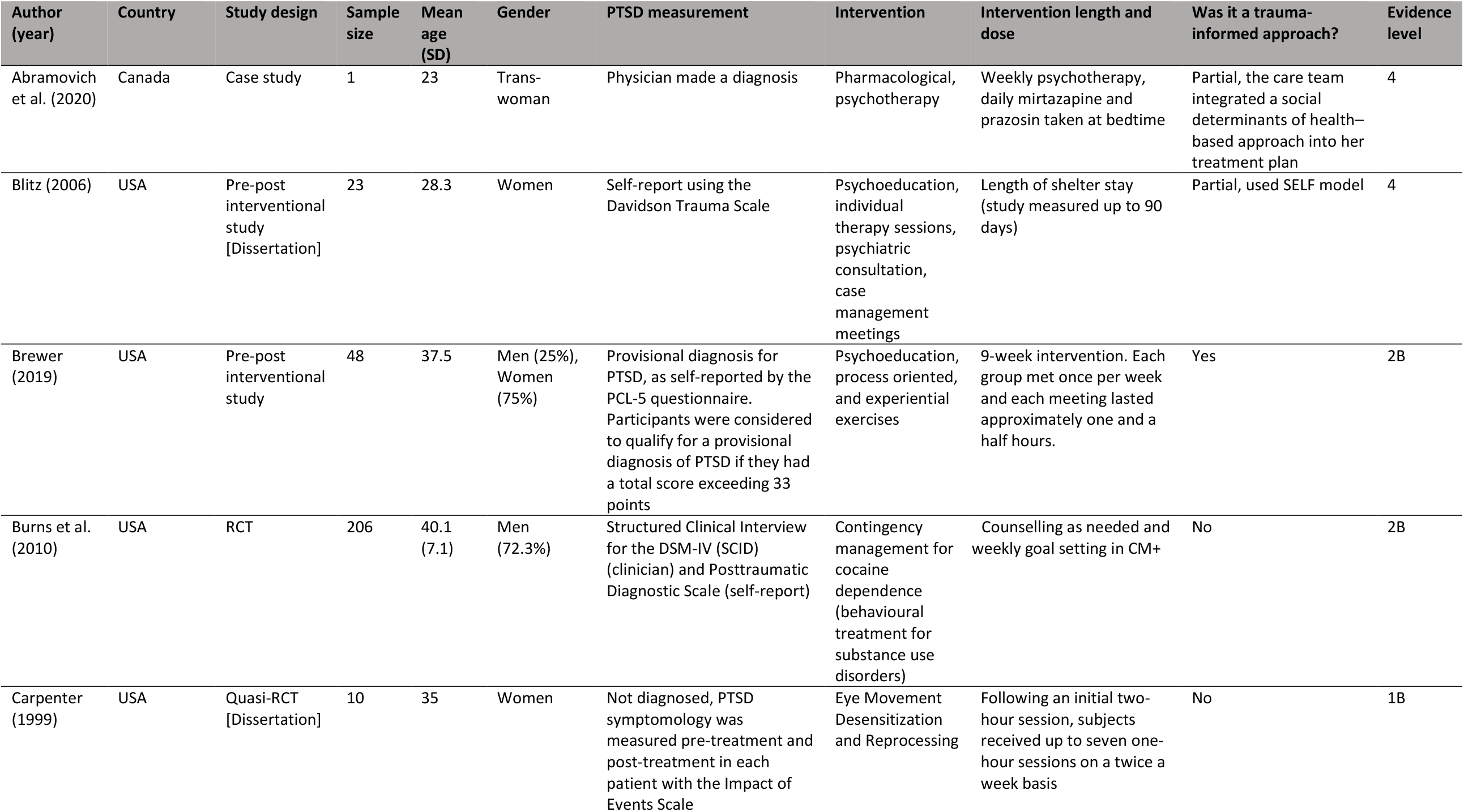
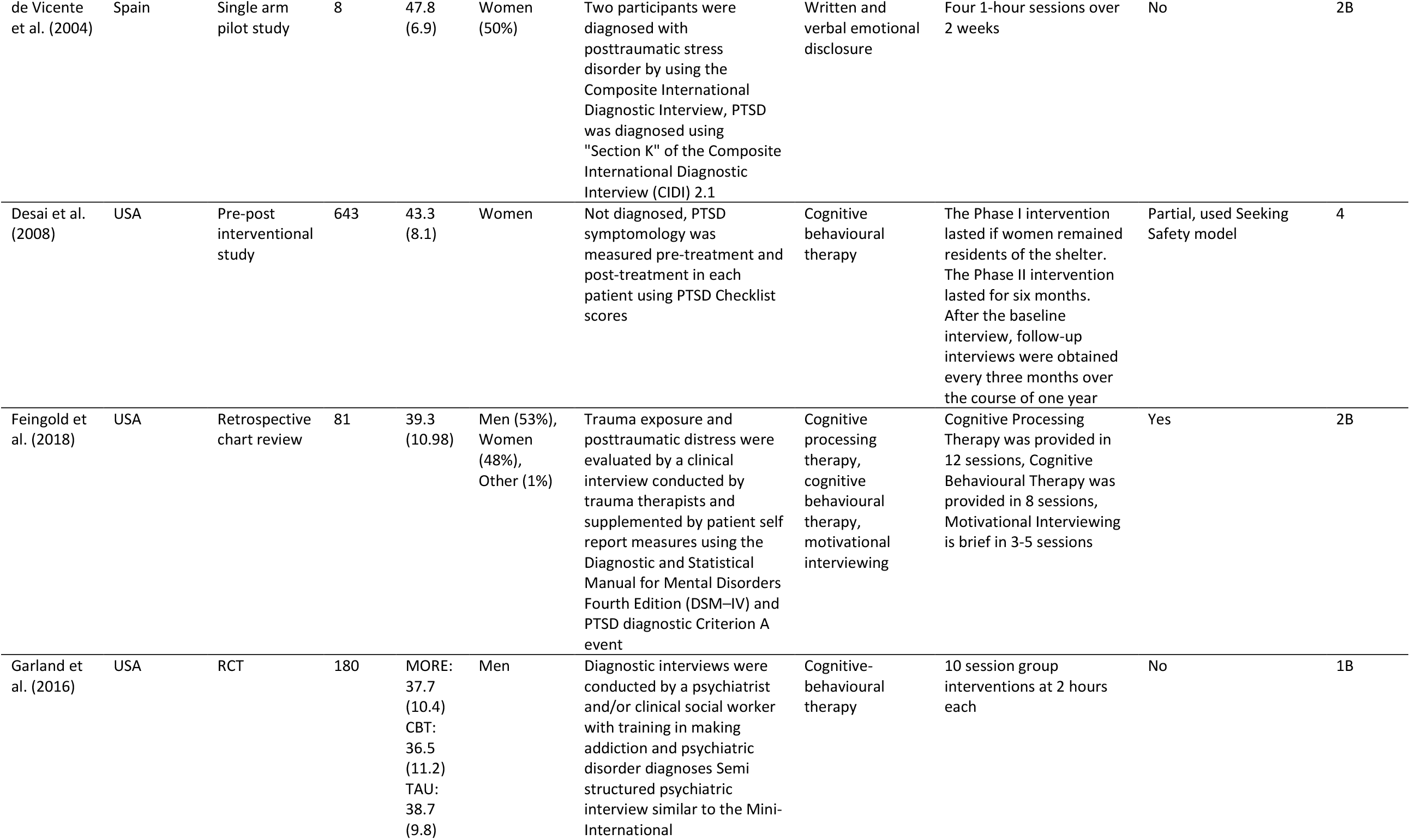
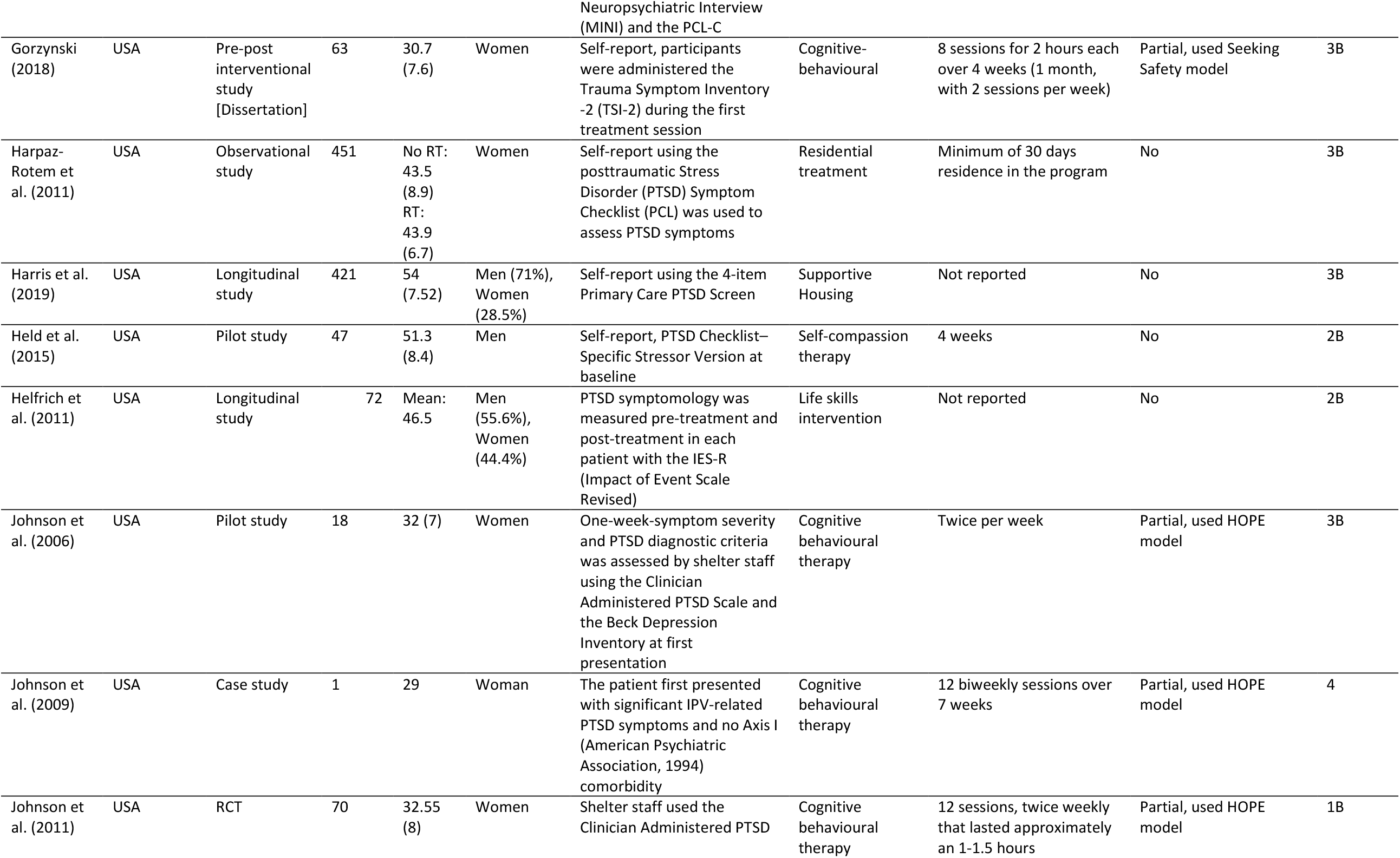
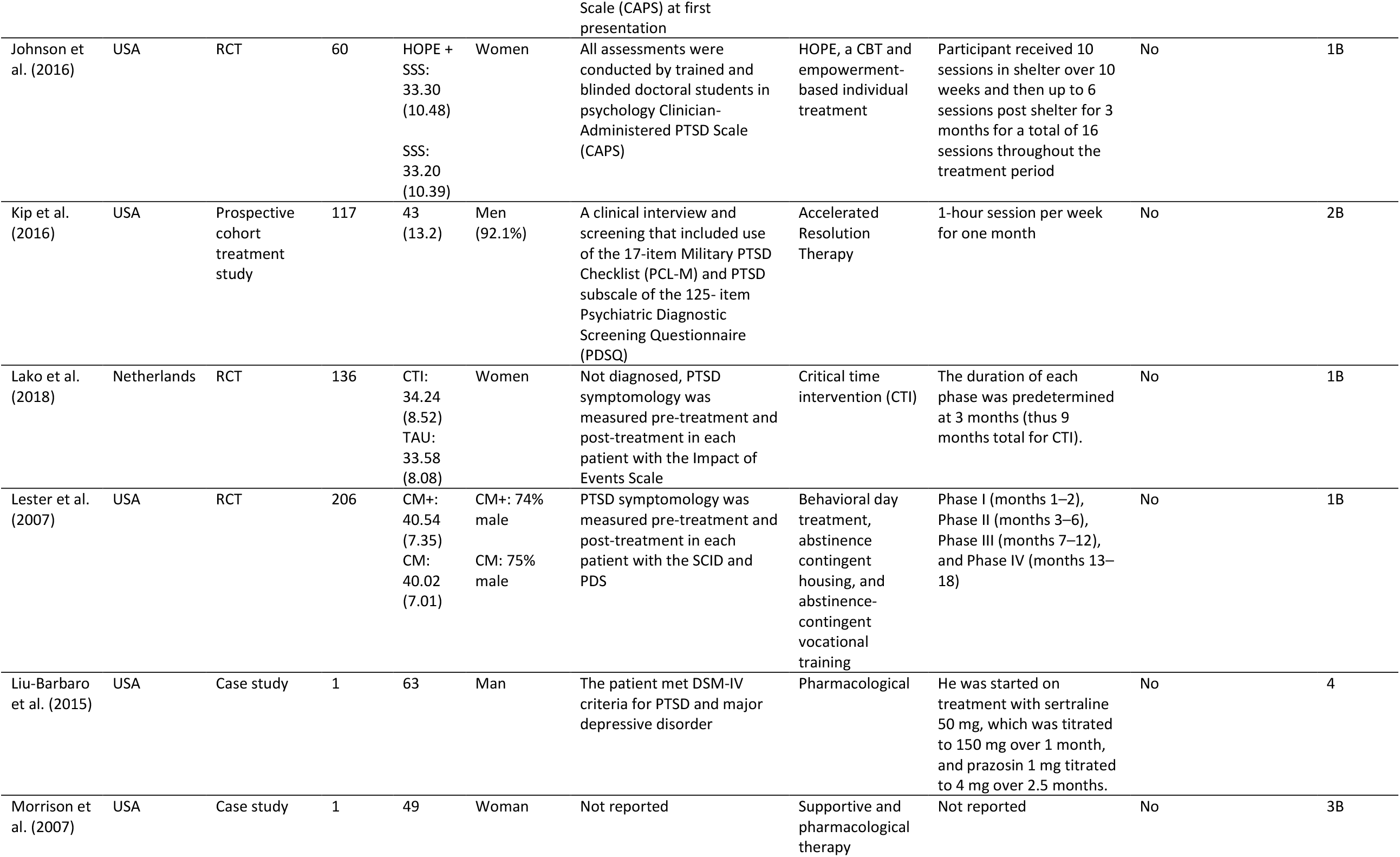
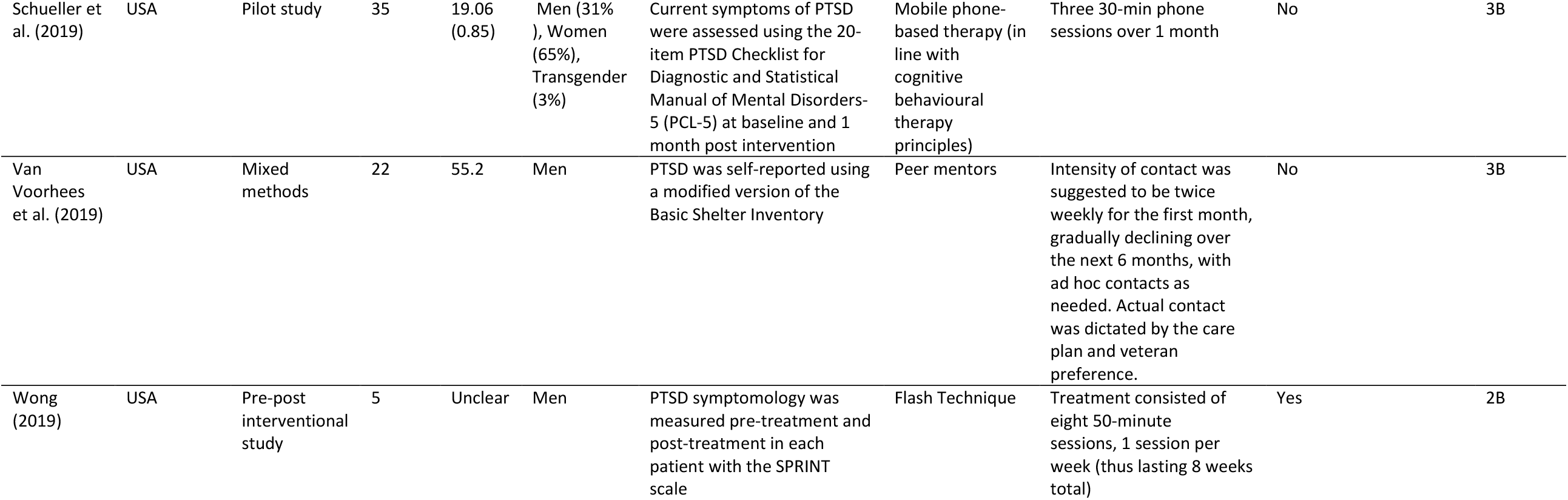
Summary of study characteristics

| Author (year) | Country | Study design | Sample size | Mean age (SD) | Gender | PTSD measurement | Intervention | Intervention length and dose | Was it a trauma-informed approach? | Evidence level |
| --- | --- | --- | --- | --- | --- | --- | --- | --- | --- | --- |
| Abramovich et al. (2020) | Canada | Case study | 1 | 23 | Trans-woman | Physician made a diagnosis | Pharmacological, psychotherapy | Weekly psychotherapy, daily mirtazapine and prazosin taken at bedtime | Partial, the care team integrated a social determinants of health–based approach into her treatment plan | 4 |
| Blitz (2006) | USA | Pre-post interventional study [Dissertation] | 23 | 28.3 | Women | Self-report using the Davidson Trauma Scale | Psychoeducation, individual therapy sessions, psychiatric consultation, case management meetings | Length of shelter stay (study measured up to 90 days) | Partial, used SELF model | 4 |
| Brewer (2019) | USA | Pre-post interventional study | 48 | 37.5 | Men (25%), Women (75%) | Provisional diagnosis for PTSD, as self-reported by the PCL-5 questionnaire. Participants were considered to qualify for a provisional diagnosis of PTSD if they had a total score exceeding 33 points | Psychoeducation, process oriented, and experiential exercises | 9-week intervention. Each group met once per week and each meeting lasted approximately one and a half hours. | Yes | 2B |
| Burns et al. (2010) | USA | RCT | 206 | 40.1 (7.1) | Men (72.3%) | Structured Clinical Interview for the DSM-IV (SCID) (clinician) and Posttraumatic Diagnostic Scale (self-report) | Contingency management for cocaine dependence (behavioural treatment for substance use disorders) | Counselling as needed and weekly goal setting in CM+ | No | 2B |
| Carpenter (1999) | USA | Quasi-RCT [Dissertation] | 10 | 35 | Women | Not diagnosed, PTSD symptomology was measured pre-treatment and post-treatment in each patient with the Impact of Events Scale | Eye Movement Desensitization and Reprocessing | Following an initial two-hour session, subjects received up to seven one-hour sessions on a twice a week basis | No | 1B |
| de Vicente et al. (2004) | Spain | Single arm pilot study | 8 | 47.8 (6.9) | Women (50%) | Two participants were diagnosed with posttraumatic stress disorder by using the Composite International Diagnostic Interview, PTSD was diagnosed using "Section K" of the Composite International Diagnostic Interview (CIDI) 2.1 | Written and verbal emotional disclosure | Four 1-hour sessions over 2 weeks | No | 2B |
| Desai et al. (2008) | USA | Pre-post interventional study | 643 | 43.3 (8.1) | Women | Not diagnosed, PTSD symptomology was measured pre-treatment and post-treatment in each patient using PTSD Checklist scores | Cognitive behavioural therapy | The Phase I intervention lasted if women remained residents of the shelter. The Phase II intervention lasted for six months. After the baseline interview, follow-up interviews were obtained every three months over the course of one year | Partial, used Seeking Safety model | 4 |
| Feingold et al. (2018) | USA | Retrospective chart review | 81 | 39.3 (10.98) | Men (53%), Women (48%), Other (1%) | Trauma exposure and posttraumatic distress were evaluated by a clinical interview conducted by trauma therapists and supplemented by patient self report measures using the Diagnostic and Statistical Manual for Mental Disorders Fourth Edition (DSM-IV) and PTSD diagnostic Criterion A event | Cognitive processing therapy, cognitive behavioural therapy, motivational interviewing | Cognitive Processing Therapy was provided in 12 sessions, Cognitive Behavioural Therapy was provided in 8 sessions, Motivational Interviewing is brief in 3-5 sessions | Yes | 2B |
| Garland et al. (2016) | USA | RCT | 180 | MORE: 37.7 (10.4)<br>CBT: 36.5 (11.2)<br>TAU: 38.7 (9.8) | Men | Diagnostic interviews were conducted by a psychiatrist and/or clinical social worker with training in making addiction and psychiatric disorder diagnoses Semi structured psychiatric interview similar to the Mini-International | Cognitive-behavioural therapy | 10 session group interventions at 2 hours each | No | 1B |

|  |  |  |  |  |  | Neuropsychiatric Interview (MINI) and the PCL-C |  |  |  |  |
| --- | --- | --- | --- | --- | --- | --- | --- | --- | --- | --- |
| Gorzynski (2018) | USA | Pre-post interventional study [Dissertation] | 63 | 30.7 (7.6) | Women | Self-report, participants were administered the Trauma Symptom Inventory -2 (TSI-2) during the first treatment session | Cognitive-behavioural | 8 sessions for 2 hours each over 4 weeks (1 month, with 2 sessions per week) | Partial, used Seeking Safety model | 3B |
| Harpaz-Rotem et al. (2011) | USA | Observational study | 451 | No RT: 43.5 (8.9)<br>RT: 43.9 (6.7) | Women | Self-report using the posttraumatic Stress Disorder (PTSD) Symptom Checklist (PCL) was used to assess PTSD symptoms | Residential treatment | Minimum of 30 days residence in the program | No | 3B |
| Harris et al. (2019) | USA | Longitudinal study | 421 | 54 (7.52) | Men (71%), Women (28.5%) | Self-report using the 4-item Primary Care PTSD Screen | Supportive Housing | Not reported | No | 3B |
| Held et al. (2015) | USA | Pilot study | 47 | 51.3 (8.4) | Men | Self-report, PTSD Checklist–Specific Stressor Version at baseline | Self-compassion therapy | 4 weeks | No | 2B |
| Helfrich et al. (2011) | USA | Longitudinal study | 72 | Mean: 46.5 | Men (55.6%), Women (44.4%) | PTSD symptomology was measured pre-treatment and post-treatment in each patient with the IES-R (Impact of Event Scale Revised) | Life skills intervention | Not reported | No | 2B |
| Johnson et al. (2006) | USA | Pilot study | 18 | 32 (7) | Women | One-week-symptom severity and PTSD diagnostic criteria was assessed by shelter staff using the Clinician Administered PTSD Scale and the Beck Depression Inventory at first presentation | Cognitive behavioural therapy | Twice per week | Partial, used HOPE model | 3B |
| Johnson et al. (2009) | USA | Case study | 1 | 29 | Woman | The patient first presented with significant IPV-related PTSD symptoms and no Axis I (American Psychiatric Association, 1994) comorbidity | Cognitive behavioural therapy | 12 biweekly sessions over 7 weeks | Partial, used HOPE model | 4 |
| Johnson et al. (2011) | USA | RCT | 70 | 32.55 (8) | Women | Shelter staff used the Clinician Administered PTSD | Cognitive behavioural therapy | 12 sessions, twice weekly that lasted approximately an 1-1.5 hours | Partial, used HOPE model | 1B |

|  |  |  |  |  |  | Scale (CAPS) at first presentation |  |  |  |  |
| --- | --- | --- | --- | --- | --- | --- | --- | --- | --- | --- |
| Johnson et al. (2016) | USA | RCT | 60 | HOPE + SSS: 33.30 (10.48)<br>SSS: 33.20 (10.39) | Women | All assessments were conducted by trained and blinded doctoral students in psychology Clinician-Administered PTSD Scale (CAPS) | HOPE, a CBT and empowerment-based individual treatment | Participant received 10 sessions in shelter over 10 weeks and then up to 6 sessions post shelter for 3 months for a total of 16 sessions throughout the treatment period | No | 1B |
| Kip et al. (2016) | USA | Prospective cohort treatment study | 117 | 43 (13.2) | Men (92.1%) | A clinical interview and screening that included use of the 17-item Military PTSD Checklist (PCL-M) and PTSD subscale of the 125- item Psychiatric Diagnostic Screening Questionnaire (PDSQ) | Accelerated Resolution Therapy | 1-hour session per week for one month | No | 2B |
| Lako et al. (2018) | Netherlands | RCT | 136 | CTI: 34.24 (8.52)<br>TAU: 33.58 (8.08) | Women | Not diagnosed, PTSD symptomology was measured pre-treatment and post-treatment in each patient with the Impact of Events Scale | Critical time intervention (CTI) | The duration of each phase was predetermined at 3 months (thus 9 months total for CTI). | No | 1B |
| Lester et al. (2007) | USA | RCT | 206 | CM+: 40.54 (7.35)<br>CM: 40.02 (7.01) | CM+: 74% male<br>CM: 75% male | PTSD symptomology was measured pre-treatment and post-treatment in each patient with the SCID and PDS | Behavioral day treatment, abstinence contingent housing, and abstinence-contingent vocational training | Phase I (months 1–2), Phase II (months 3–6), Phase III (months 7–12), and Phase IV (months 13–18) | No | 1B |
| Liu-Barbaro et al. (2015) | USA | Case study | 1 | 63 | Man | The patient met DSM-IV criteria for PTSD and major depressive disorder | Pharmacological | He was started on treatment with sertraline 50 mg, which was titrated to 150 mg over 1 month, and prazosin 1 mg titrated to 4 mg over 2.5 months. | No | 4 |
| Morrison et al. (2007) | USA | Case study | 1 | 49 | Woman | Not reported | Supportive and pharmacological therapy | Not reported | No | 3B |
| Schueller et al. (2019) | USA | Pilot study | 35 | 19.06 (0.85) | Men (31%), Women (65%), Transgender (3%) | Current symptoms of PTSD were assessed using the 20-item PTSD Checklist for Diagnostic and Statistical Manual of Mental Disorders-5 (PCL-5) at baseline and 1 month post intervention | Mobile phone-based therapy (in line with cognitive behavioural therapy principles) | Three 30-min phone sessions over 1 month | No | 3B |
| Van Voorhees et al. (2019) | USA | Mixed methods | 22 | 55.2 | Men | PTSD was self-reported using a modified version of the Basic Shelter Inventory | Peer mentors | Intensity of contact was suggested to be twice weekly for the first month, gradually declining over the next 6 months, with ad hoc contacts as needed. Actual contact was dictated by the care plan and veteran preference. | No | 3B |
| Wong (2019) | USA | Pre-post interventional study | 5 | Unclear | Men | PTSD symptomology was measured pre-treatment and post-treatment in each patient with the SPRINT scale | Flash Technique | Treatment consisted of eight 50-minute sessions, 1 session per week (thus lasting 8 weeks total) | Yes | 2B |

### Research question 1: What interventions are described in the literature for the treatment of PTSD and cPTSD in individuals who are vulnerably housed?

#### Trauma focused psychotherapies

We identified four studies that used an intervention where participants were asked to recall details of the traumatic events. One non-randomized study evaluated the effectiveness of using eye movement desensitization and reprocessing (EMDR) in 10 women living in a battered women’s shelter in Michigan.[31] Both the five women who received EMDR and the five women who received only the standard shelter program experienced significant reductions in PTSD symptoms although the reduction was greater in the EMDR group. One case series of five men attending a trauma group in a Californian Homeless Shelter study used the Flash Technique [32] which is part of the preparatory phase of EMDR treatment. Another observational study evaluated the feasibility of using accelerated resolution therapy [33] in a cohort of 23 homeless veterans compared to 94 community living veterans in Tampa, Florida. Accelerated resolution therapy involves brief recall of traumatic events, imagery rescripting and some elements of EMDR. About half of the homeless group completed treatment compared to 80% of community veterans. The reasons for non-completion in the homeless were moving away from the shelter, conflict with work, and other life circumstances. The authors comment that therapies for the homeless need to be brief and need to take into account a range of comorbidities and significant life challenges. They also commented on the difficulties of finding a private quiet setting which isn’t always available in a homeless shelter. De Vicente et al[34] described a case series of 8 homeless people, two of whom were diagnosed with PTSD, attending a day centre in Madrid, Spain for the homeless. They received an emotional disclosure protocol where participants were asked to write or speak about their thoughts and feelings associated with their trauma.

### Non-trauma psychotherapies

Johnson et al. evaluated a cognitive-behavioral therapy (CBT) and empowerment-based individual treatment called Health to Overcome PTSD through Empowerment (HOPE). They reported a case study, pilot study, and two RCTs in women with PTSD or subthreshold PTSD from “battered women’s shelters” comparing HOPE with “standard shelter treatment”.[35–38] The HOPE therapy focused on addressing PTSD resulting from intimate partner violence, while using many traditional components of CBT for PTSD (for example cognitive-restructuring, skill building) with a focus on stabilization, safety and empowerment. In the first RCT of 70 people, they found no difference in PTSD outcomes with one in four participants attending all 12 HOPE sessions and about two-thirds attending five or more sessions. Participants were excluded if they were psychotic, suicidal, had been diagnosed with bipolar disorder or had any change in psychotropic medication in the previous month. In the second RCT of 60 women in shelters, the treatment was continued after the women had left the shelter but again did not find a significant difference in PTSD outcomes.

Two observational studies aimed to evaluate the effectiveness of a CBT based group therapy, Seeking Safety in homeless women.[39,40] Seeking Safety is a manualized CBT intervention consisting of 25 individual modules that address issues of safe behaviors and relationships, life skills, and relapse prevention. In the 2008 observational cohort study, 91 female US veterans who were vulnerably housed were offered Seeking Safety and were compared to a historical cohort of 359 women who did not receive Seeking Safety. It is unclear how many of the women had PTSD but there was a significant decrease in PTSD symptoms over the whole group although the differences were small. The Seeking Safety group significantly increased their drug use. In the 2017 pretest post-test interventional study, 63 homeless women in an in-patient residential setting who had been exposed to trauma and had substance use disorders were offered an abbreviated version of Seeking Safety. The abbreviated version was developed in response to criticism that the full version was too long, required high levels of participant commitment, and was unsuitable for “transient” populations. The authors found that after eight two-hour sessions over four weeks there was a significant improvement in perceived self-efficacy in the two-thirds of the sample who completed a minimum of six out of eight sessions. The Veterans Affairs Clinical Practice Guidelines concludes that there is insufficient evidence to recommend for or against Seeking Safety for treatment of PTSD in the general population.

Feingold [41] reported an observational cohort study of cognitive processing therapy, CBT and motivational interviewing in adults within a US jail diversion program. About a third of their population of 81 participants were vulnerably housed and 62% were diagnosed with PTSD. The main finding was that about half of the sample did not complete their treatment. Increased completion rates were associated with emergency therapy sessions and the authors comment on the need for flexibility in delivering care to this population. In those who did complete their treatment there was a significant decrease in PTSD symptoms.

Schueller [42] reported a single group pilot study evaluating the feasibility and acceptability of a mobile phone-based therapy that used an app designed to help provide coping skills to 35 young adults (18-24 years) recruited from homeless shelters in Chicago. Participants received a mobile phone, a data plan, the app and one month of support from a coach consisting of up to three brief sessions. Just over half of the participants completed all three coaching sessions but there were no significant changes in clinical outcomes.

#### Contingency management

In one unblinded RCT, 187 homeless people from Alabama, dependent on cocaine, were randomized to contingency management or contingency management plus. At the start of the study 21 participants had a diagnosis of PTSD. Housing was dependent on negative urine screens for cocaine. There did not appear to be any difference in outcomes between the two groups.[43,44]

#### Educational interventions

One RCT examined the effectiveness of a critical time intervention for abused women who were transitioning from women’s shelters to community living.[45] Critical time intervention is a time-limited, strengths-based intervention designed to support vulnerable people during transitions in their lives. This study of 136 Dutch women found a significant decrease in PTSD symptoms after nine months which was mainly due to a reduction in symptoms in non-Dutch speaking women. Another pilot study evaluated the effect of self-compassion therapy compared to stress inoculation on trauma-related guilt and PTSD severity.[46] The authors randomized 47 US veterans living in transitional housing to receive workbooks on the different therapies. Symptoms of PTSD did not significantly differ between the two groups after treatment. Twenty of the participants did not complete the workbooks. One qualitative study of 22 homeless US veterans hypothesised that feeling disconnected was associated with poor outcomes in those with PTSD.[47] A pretest and post-test study of 8 adults looked to evaluate the treatment effects of Building Bridges, a group-based psychotherapy adapted for parents living in homeless shelters in San Francisco.[48] Building Bridges consisted of nine 90 minute groups that included psychoeducation, mindfulness and experiential exercises. There was no significant impact on PTSD symptoms. The authors comment on the difficulty of providing groups in a shelter setting because of staff turnover and restrictions, such as length of stay, at different sites. Another pretest post-test study examined the impact of a life skills group and individual program on 72 individuals who had been recently homeless living in emergency housing.[49] Eight out of ten participants reported a history of abuse. Symptoms of PTSD significantly decreased after the intervention. The life skills intervention consisted of one or more modules which focused on food management, money management, room and self-care management and safe community participation.

### Housing interventions

We identified three studies that used housing and shelter as a way to provide therapy.[50–52] One study used a shelter as a restorative milieu that integrated trauma recovery and social justice empowerment for Black and Latino women in New York.[50] This pretest post-test descriptive and qualitative study of about 20 women found a reduction in PTSD symptoms over time and an appreciation by the residents that the shelter milieu felt safe and nurturing. Another study focused on providing supportive housing to 421 homeless adults in Los Angeles.[52] This descriptive pretest and post-test study found that about half the sample had probable PTSD at baseline which reduced to 40% at three, six and 12 months. A cohort study of homeless US female veterans compared 234 who received at least 30 days of residential treatment with 217 who received less than 30 days. The study was able to follow up about half the participants after one year. The authors found a small but significant improvement in PTSD symptoms in those who received residential treatment.[51]

### Pharmacotherapy

There were three single person case studies describing successful pharmacological treatments.[53–55] In one study, a 23-year-old transgender refugee woman in an emergency shelter in Canada was prescribed mirtazapine 60mg for PTSD and insomnia, and prazosin 6mg for nightmares.[53] Liu-Barbaro and Stein[54] describe treating a 63-year-old Ethiopian man living in a homeless shelter with a major depressive disorder and PTSD with dissociative symptoms. He was treated successfully with sertraline 150mg and prazosin 4mg. The authors note that this patient had suffered for years before being diagnosed and treated.[54] Finally, the last study reported a “composite case” of a 49-year-old woman with PTSD prescribed sertraline 150mg and supportive therapy.[55]

### Research question 2: Are there any interventions for treatment of PTSD and cPTSD in the vulnerably housed which are described as trauma-informed, and how is this operationalized?

We identified three studies that explicitly stated using a trauma-informed approach to deliver treatment.[32,41,48] Two studies did not clearly describe how they operationalized their trauma-informed approach.[32,41] One study investigating the Building Bridges intervention described using a trauma-informed approach based on the work by Guarino et al.[56] As a result Building Bridges is based on understanding how trauma affects parents’ and children’s cognitions, emotions and behaviors. The authors stressed that emotional safety, and the desire to feel heard, protected, comforted, and understood is an important aspect in treating populations who have experienced trauma.

There were 7 studies that did not explicitly state using a trauma-informed approach; however they did describe components of their treatment that align with the concepts of a trauma-informed care.[35–39,50,53,57] The HOPE intervention focused on the impact of present trauma of interpersonal violence and did not focus on trauma from the past. The research group investigating HOPE used Herman’s[58] multistage model of recovery which addresses the treatment needs of battered women and incorporates three stages of recovery: (a) establishing safety, self-care, and protection, (b) remembrance and mourning, and (c) reconnection. The authors also note the importance of flexibility and being available when people are ready to engage.[59] Two studies used Seeking Safety[39,57] which they describe as based on five central ideas: “1) Safety as the priority of this first stage treatment; 2) integrated treatment of PTSD and substance abuse; 3) a focus on ideals; 4) four content areas: cognitive, behavioral, interpersonal, and case management; and 5) attention to therapeutic processes.” The goals of the treatment are to create an empathetic approach where the clients “own” the trauma, to provide education, to validate the connection between trauma and substance abuse, and to offer safe coping skills to manage the symptoms, impulses, and emotions that often come with these co-occurring disorders. One case study integrated a social determinants of health based approach into the treatment plan.[53]

## Discussion

Despite the high prevalence of PTSD in the vulnerably housed there is little evidence of what is effective treatment. Figure 2 outlines how the 26 studies identified map onto the four components of trauma-informed care. The four trauma-informed interventions were small case series, and the non-trauma focused therapies were generally ineffective which is consistent with the broader literature. A novel intervention in this population is using housing as an intervention but the impact on PTSD symptoms was relatively modest and the strength of evidence low. A systematic review of permanent supportive housing for homeless individuals found no impact on psychiatric symptoms although there was greater housing stability compared to usual care.[60] What is notable is the difficulty of keeping people in treatment with most studies reporting low engagement with treatment or low follow-up rates. Few studies described what proportion of people agreed to participate in the treatments but the small numbers in most studies suggest that engagement in treatment is difficult.

**Figure 2.**
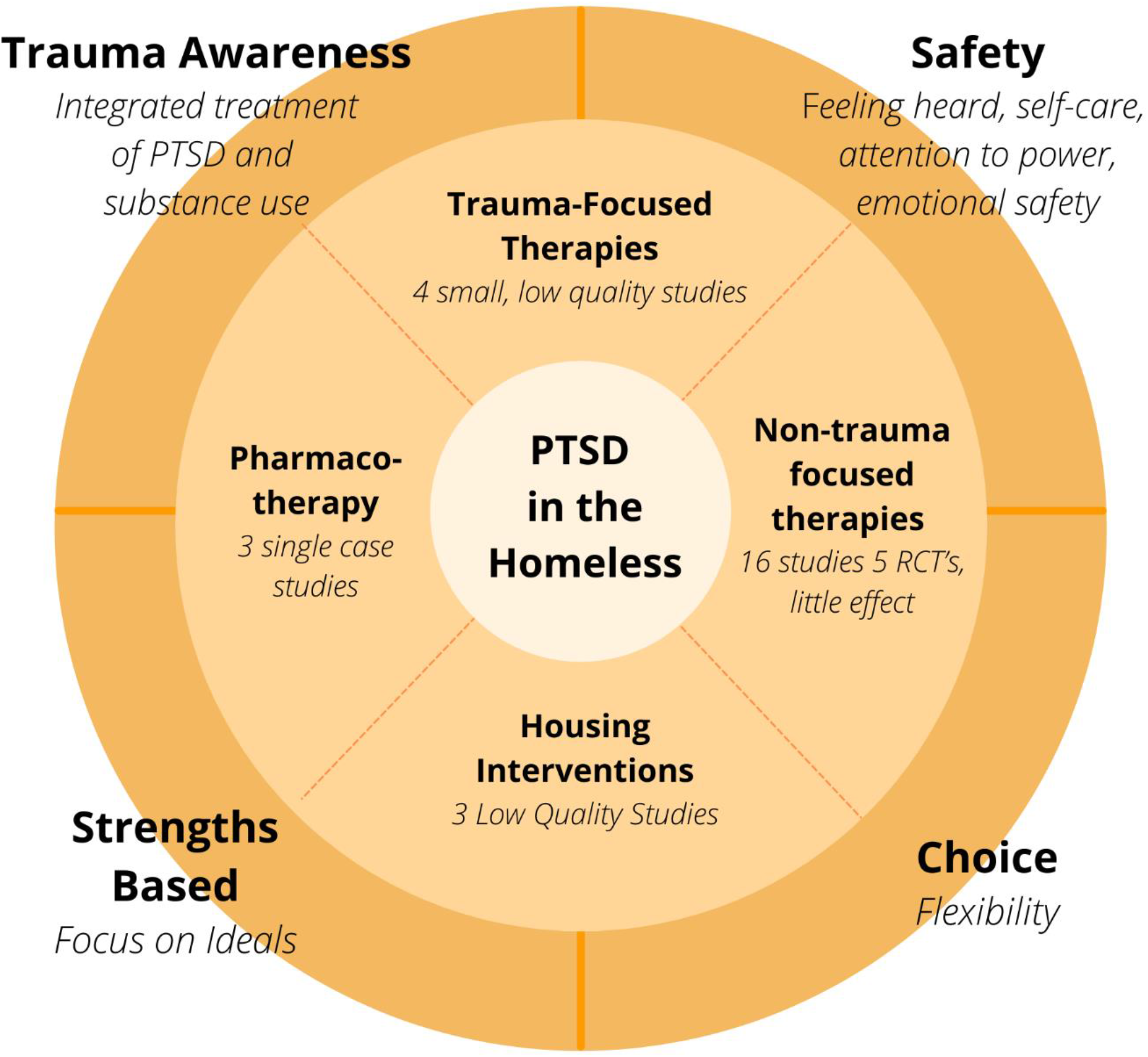
Summary of findings. 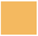 Trauma informed care (from Hopper) described in the scoping review (Outer Circle) 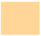 Types of intervention found in the scoping review (Middle Circle)

We found three studies that explicitly stated using a trauma-informed approach, while seven described a method that incorporated all or some components of a trauma-informed care approach. Although guidelines and organizations recognize the need for a trauma-informed approach,[20,21,61,62] it is unclear how this translates into action or what the most important components are. This review highlights the need to operationalize trauma-informed care and to identify the necessary and most important components in the vulnerably housed. Another area that is unexplored is whether what people with lived experience consider to be trauma-informed care is different to what providers would consider. The impact of an intervention that is trauma-informed is most likely to be seen in increased rates of engagement and completion of treatment.

Our review has some important limitations to note. Firstly, the interventions to treat PTSD in the vulnerably housed are primarily studied in women and US Veterans. Similarly, many studies did not mention any ethnic or cultural factors to consider when providing treatment for the vulnerably housed. Second, is that there is a lack of qualitative and experiential data to illustrate any other meaningful changes that may have occurred during treatment.

Lastly, there is low methodological quality and reporting of studies. Although a formal quality assessment was not performed, the levels of evidence were rated for each study and most studies were rated lower quality with small sample sizes or low rates of follow-up. Any conclusions made from these studies should be interpreted with caution.

## Conclusion

There is currently little evidence on how trauma-informed care for PTSD in the vulnerably housed should be delivered and whether it is effective. Poor quality trials make interpretation of acceptability, feasibility and effectiveness difficult. Evaluation of interventions should be expanded to include not only symptom improvement, but experiential data informed by the engagement of patients as partners. Measures of symptom severity alone often do not provide a complete picture of the patient experience, excluding factors that may be important such as increased understanding of their illness, skill building, coping and wellbeing.

This review has also highlighted the need for pragmatic trial designs instead of “one-size fits all” interventions and delivery approaches. The development of a trauma-informed care strategy should be adaptable to multiple cultural or geographic situations to ensure that healthcare providers are able to deliver meaningful, evidence-based care and no individuals are “left behind”. Any guidance should include recommendations for implementation or adaptation to ensure fidelity for comparison of effectiveness while allowing for flexibility in delivery.

Managing PTSD and complex PTSD can be challenging for many service providers in a population where appropriate treatment approaches are poorly understood, under-researched and lack a patient-oriented perspective. This scoping review has identified several gaps in providing trauma-informed care to a vulnerable population. There is a need to conduct well-designed trials with mixed-methods approaches, focusing on trauma-informed care principles to improve treatments for the vulnerably housed experiencing or at high risk of developing mental health issues. Healthcare providers and policymakers need more guidance on working with the vulnerably housed, including on how to collect data and outcomes specific to trauma-informed care.

## Data Availability

All data relevant to the study are included in the article or uploaded as supplemental information.

## Contributors

This research was conceptualized by SH, while AB and NEE contributed to the study design. Search Strategy and searches were completed by AH. While title, abstract and full screenings, and data extraction were carried out by AB, KC, MK under guidance from SH. Manuscript was drafted by AB, NEE and SH. Critical review of manuscript was undertaken by all authors. All authors approved the final manuscript.

## Data sharing statement

All data relevant to the study are included in the article or uploaded as supplemental information.

