## Supplemental File 1 for "Interventions to treat post-traumatic stress disorder (PTSD) in vulnerably housed populations and trauma-informed care: A scoping review"

### Appendix 1 - Final literature search strategies

Limits: none

Searches run on March 26, 2020

Databases:

- Medline (medall)
- Embase (emczd)
- APA PsycInfo (psyh)
- CINAHL
- Cochrane Library (via Wiley platform)
- WebOfScience
- PtsdPubs

**Embase, Ovid MEDLINE(R), APA PsycInfo**

Search history sorted by search number ascending

| # | Searches | Results |
|---|---|---|
| 1 | exp Homeless Persons/ | 10564 |
| 2 | homeless*.ti,ab,kf. | 33339 |
| 3 | ((street* or no home or no homes) adj3 (people or youth or person? or population? or individual? or m?n or wom?n or adult?)).ti,ab,kf. | 3254 |
| 4 | ((hous* or home or homes or shelter* or hostel? or accomodation? or dwelling?) adj3 (insecur* or instabil* or stabil* or stable or unstabl* or temporar* or marginal* or precarious* or inadequate*)).ti,ab,kf. | 12953 |
| 5 | housing outcome*.ti,ab,kf. | 249 |
| 6 | (vulnerabl* adj2 hous*).ti,ab,kf. | 564 |
| 7 | (no housing or no fixed address or squatter* or evict*).ti,ab,kf. | 3651 |
| 8 | or/1-7 | 52487 |
| 9 | exp housing/ | 68128 |
| 10 | housing.ti,ab,kf. | 80001 |
| 11 | or/9-10 | 117889 |
| 12 | vulnerable populations/ | 23575 |
| 13 | exp poverty/ | 97990 |
| 14 | ((low adj3 (income* or revenu*)) or poor or poverty or vulnerabl*).ti,ab,kf. | 1851735 |
| 15 | ((marginal or precarious* or disadvantag* or "at risk") adj3 (people or youth or person? or population? or individual? or m?n or wom?n or adult?)).ti,ab,kf. | 120846 |
| 16 | or/12-15 | 2005185 |
| 17 | and/11,16 | 17080 |

| | | |
|---|---|---|
| 18 | or/8,17 | 65715 |
| 19 | "trauma and stressor related disorders"/ | 48773 |
| 20 | exp stress disorders, traumatic/ | 94387 |
| 21 | (post-traumatic or posttraumatic or ptsd or traumatic disorder*).ti,ab,kf. | 203057 |
| 22 | ((posttrauma* or post-trauma*) adj3 (stress* or disorder* or psych* or symptom*)).ti,ab,kf. | 115064 |
| 23 | (acute stress disorder* or combat disorder* or war neuros*).ti,ab,kf. | 2822 |
| 24 | (trauma-informed or trauma-focus*).ti,ab,kf. | 6927 |
| 25 | (trauma* adj3 (care or treatment* or therap* or intervention*)).ti,ab,kf. | 58728 |
| 26 | or/19-25 | 281197 |
| 27 | and/18,26 | 1582 |
| 28 | 27 use medall | 473 |
| 29 | exp homeless person/ | 10564 |
| 30 | homeless*.ti,ab,kw. | 33547 |
| 31 | ((street* or no home or no homes) adj3 (people or youth or person? or population? or individual? or m?n or wom?n or adult?)).ti,ab,kw. | 3262 |
| 32 | ((hous* or home or homes or shelter* or hostel? or accomodation? or dwelling?) adj3 (insecur* or instabil* or stabil* or stable or unstabl* or temporar* or marginal* or precarious* or inadequate*)).ti,ab,kw. | 12990 |
| 33 | housing outcome*.ti,ab,kw. | 249 |
| 34 | (vulnerabl* adj2 hous*).ti,ab,kw. | 578 |
| 35 | (no housing or no fixed address or squatter* or evict*).ti,ab,kw. | 3669 |
| 36 | or/29-35 | 52686 |
| 37 | housing/ | 49217 |
| 38 | real estate/ | 231 |
| 39 | housing.ti,ab,kw. | 80421 |
| 40 | or/37-39 | 103700 |
| 41 | vulnerable population/ | 26323 |
| 42 | poverty/ | 92527 |
| 43 | exp lowest income group/ | 27543 |
| 44 | ((low adj3 (income* or revenu*)) or poor or poverty or vulnerabl*).ti,ab,kw. | 1853972 |
| 45 | ((marginal or precarious* or disadvantag* or "at risk") adj3 (people or youth or person? or population? or individual? or m?n or wom?n or adult?)).ti,ab,kw. | 120465 |
| 46 | or/41-45 | 2010908 |
| 47 | and/40,46 | 16486 |
| 48 | or/36,47 | 65390 |
| 49 | posttraumatic stress disorder/ | 122887 |
| 50 | psychotrauma/ | 9075 |
| 51 | (post-traumatic or posttraumatic or ptsd or traumatic disorder*).ti,ab,kw. | 204775 |

| | | |
|---|---|---|
| 52 | ((posttrauma* or post-trauma*) adj3 (stress* or disorder* or psych* or symptom*)).ti,ab,kw. | 116228 |
| 53 | (acute stress disorder* or combat disorder* or war neuros*).ti,ab,kw. | 2936 |
| 54 | (trauma-informed or trauma-focus*).ti,ab,kw. | 6948 |
| 55 | (trauma* adj3 (care or treatment* or therap* or intervention*)).ti,ab,kw. | 59229 |
| 56 | or/49-55 | 287064 |
| 57 | and/48,56 | 1606 |
| 58 | 57 use emczd | 679 |
| 59 | exp homeless/ | 7407 |
| 60 | homeless*.ti,ab. | 33202 |
| 61 | ((street* or no home or no homes) adj3 (people or youth or person? or population? or individual? or m?n or wom?n or adult?)).ti,ab. | 3245 |
| 62 | ((hous* or home or homes or shelter* or hostel? or accomodation? or dwelling?) adj3 (insecur* or instabil* or stabil* or stable or unstabl* or temporar* or marginal* or precarious* or inadequate*)).ti,ab. | 12920 |
| 63 | housing outcome*.ti,ab. | 249 |
| 64 | (vulnerabl* adj2 hous*).ti,ab. | 562 |
| 65 | (no housing or no fixed address or squatter* or evict*).ti,ab. | 3638 |
| 66 | or/59-65 | 50449 |
| 67 | exp housing/ | 68128 |
| 68 | exp living arrangements/ | 68050 |
| 69 | housing.ti,ab. | 79147 |
| 70 | or/67-69 | 164338 |
| 71 | at risk populations/ | 37373 |
| 72 | poverty/ | 92527 |
| 73 | runaway behavior/ | 1361 |
| 74 | lower income level/ | 8653 |
| 75 | disadvantaged/ | 17646 |
| 76 | ((low adj3 (income* or revenu*)) or poor or poverty or vulnerabl*).ti,ab. | 1849039 |
| 77 | ((marginal or precarious* or disadvantag* or "at risk") adj3 (people or youth or person? or population? or individual? or m?n or wom?n or adult?)).ti,ab. | 120201 |
| 78 | or/71-77 | 2029780 |
| 79 | and/70,78 | 23725 |
| 80 | or/66,79 | 70172 |
| 81 | "Stress and Trauma Related Disorders"/ | 13 |
| 82 | exp Posttraumatic Stress Disorder/ | 122996 |
| 83 | Acute Stress Disorder/ | 2410 |
| 84 | Post-Traumatic Stress/ | 54050 |
| 85 | combat experience/ | 2864 |
| 86 | emotional trauma/ | 24247 |
| 87 | traumatic neurosis/ | 334 |

| | | |
|---|---|---|
| 88 | stress reactions/ | 85294 |
| 89 | trauma/ | 480688 |
| 90 | (post-traumatic or posttraumatic or ptsd or traumatic disorder*).ti,ab. | 201831 |
| 91 | ((posttrauma* or post-trauma*) adj3 (stress* or disorder* or psych* or symptom*)).ti,ab. | 114265 |
| 92 | (acute stress disorder* or combat disorder* or war neuros*).ti,ab. | 2797 |
| 93 | or/81-92 | 788522 |
| 94 | exp "stress and trauma related disorders"/ | 33752 |
| 95 | exp trauma/ | 3330169 |
| 96 | or/94-95 | 3354276 |
| 97 | exp intervention/ | 103258 |
| 98 | and/96-97 | 4077 |
| 99 | (trauma-informed or trauma-focus*).ti,ab. | 6863 |
| 100 | (trauma* adj3 (care or treatment* or therap* or intervention*)).ti,ab. | 58567 |
| 101 | or/98-100 | 64494 |
| 102 | or/93,101 | 822408 |
| 103 | and/80,102 | 2442 |
| 104 | 103 use psyh | 712 |
| 105 | or/28,58,104 | 1864 |
| 106 | remove duplicates from 105 | 1209 |

**CINAHL**

| # | Query | Results |
|---|---|---|
| S7 | S5 AND S6 | 620 |
| S6 | (MH "Stress Disorders, Post-Traumatic+") OR TX ( post-traumatic OR posttraumatic OR ptsd OR "traumatic disorder*" ) OR TX ( (posttrauma* OR post-trauma*) N3 (stress* OR disorder* OR psych* OR symptom*) ) OR TX ( "acute stress disorder*" OR "combat disorder*" OR "war neuros*" ) OR TX ( trauma-informed OR trauma-focus* ) OR TX ( trauma* N3 (care OR treatment* OR therap* OR intervention*) ) | 57,274 |
| S5 | S1 OR S4 | 20,662 |
| S4 | S2 AND S3 | 5,267 |

| | | |
|---|---|---|
| S3 | ( (MH "Poverty") OR (MH "Indigent Persons") OR (MH "Vulnerability") ) OR TX ( poor OR poverty OR vulnerabl* ) OR TX ( low N3 (income* or revenu*) ) OR TX ( (marginal OR precarious* OR disadvantag* OR "at risk") N3 (people OR youth OR person? OR population? OR individual? OR m?n OR wom?n OR adult?) ) | 322,387 |
| S2 | (MH "Housing+") OR TX housing* | 23,749 |
| S1 | ( (MH "Homelessness") OR (MH "Homeless Persons") ) OR TX homeless* OR TX ( (street* OR "no home" OR "no homes") N3 (people OR youth OR person? OR population? OR individual? OR m?n OR wom?n OR adult?) ) OR TX ( (hous* OR home OR homes OR shelter* OR hostel? OR accomodation? OR dwelling?) N3 (insecur* OR instabil* OR stabil* OR stable OR unstabl* OR temporar* OR marginal* OR precarious* OR inadequate*) ) OR TX ( "housing outcome" OR "housing outcomes" OR "no housing" OR "no fixed address" OR squatter* OR evict* ) OR TX vulnerabl* N2 hous* | 16,888 |

Web of Science:

| | | |
|---|---|---|
| **Set** | **Results** | **Save History / Create AlertOpen Saved History** |
| # 7 | **655** | #6 AND #5<br>*Indexes=SCI-EXPANDED, SSCI, A&HCI, CPCI-S, CPCI-SSH, ESCI Timespan=All years* |
| # 6 | **112,391** | **TOPIC:** (post-traumatic OR posttraumatic OR ptsd OR "traumatic disorder*") *OR* **TOPIC:** ((posttrauma* OR post-trauma*) NEAR/3 (stress* OR disorder* OR psych* OR symptom*)) *OR* **TOPIC:** ("acute stress disorder*" OR "combat disorder*" OR "war neuros*") *OR* **TOPIC:** (trauma-informed OR trauma- |

| | | |
|---|---|---|
| | | focus*) *OR* **TOPIC:** (trauma* NEAR/3 (care OR treatment* OR therap* OR intervention*)) *Indexes=SCI-EXPANDED, SSCI, A&HCI, CPCI-S, CPCI-SSH, ESCI Timespan=All years* |
| # 5 | **45,160** | #4 OR #1 *Indexes=SCI-EXPANDED, SSCI, A&HCI, CPCI-S, CPCI-SSH, ESCI Timespan=All years* |
| # 4 | **13,545** | #3 AND #2 *Indexes=SCI-EXPANDED, SSCI, A&HCI, CPCI-S, CPCI-SSH, ESCI Timespan=All years* |
| # 3 | **1,147,594** | **TOPIC:** (poor OR poverty OR vulnerabl*) *OR* **TOPIC:** (low NEAR/3 (income*)) *OR* **TOPIC:** ((marginal OR precarious* OR disadvantag* OR "at risk") NEAR/3 (people OR youth OR person? OR population? OR individual? OR m?n OR wom?n OR adult?)) *Indexes=SCI-EXPANDED, SSCI, A&HCI, CPCI-S, CPCI-SSH, ESCI Timespan=All years* |
| # 2 | **100,783** | **TOPIC:** (housing*) *Indexes=SCI-EXPANDED, SSCI, A&HCI, CPCI-S, CPCI-SSH, ESCI Timespan=All years* |
| # 1 | **33,943** | **TOPIC:** (homeless* OR "housing outcome" OR "housing outcomes" OR "no housing" OR "no fixed address" OR squatter* OR evict*) *OR* **TOPIC:** ((street* OR "no home" OR "no homes") NEAR/3 (people OR youth OR person? OR population? OR individual? OR m?n OR wom?n OR adult?)) *OR* **TOPIC:** ((hous* OR home OR homes OR shelter* OR hostel? OR accomodation? OR dwelling?) NEAR/3 (insecur* OR instabil* OR stabil* OR stable OR unstabl* OR temporar* OR marginal* OR precarious* OR inadequate*)) *OR* **TOPIC:** (vulnerabl* NEAR/2 hous*) *Indexes=SCI-EXPANDED, SSCI, A&HCI, CPCI-S, CPCI-SSH, ESCI Timespan=All years* |

Cochrane Library (61 references from CENTRAL only) - no results in Cochrane Database of Systematic Reviews, although the entire library was searched.

Search Name:

Date Run: 26/03/2020 18:04:44

Comment:

ID Search Hits

#1 MeSH descriptor: [Homeless Persons] explode all trees 332

#2 (homeless* OR "no housing" OR "no fixed address" OR squatter* OR evict* OR "housing outcome*"):ti,ab,kw OR (vulnerabl* NEAR/2 hous*):ti,ab,kw OR ((street* OR "no home" OR "no homes") NEAR/3 (people OR youth OR person? OR population? OR individual? OR m?n OR wom?n OR adult?)):ti,ab,kw OR ((hous* OR home OR homes OR shelter* OR hostel? OR accomodation? OR dwelling?) NEAR/3 (insecur* OR instabil* OR stabil* OR stable OR unstabl* OR temporar* OR marginal* OR precarious* OR inadequate*)):ti,ab,kw (Word variations have been searched) 1258

#3 #1 OR #2 1258

#4 MeSH descriptor: [Housing] explode all trees 393

#5 (housing*):ti,ab,kw 1497

#6 #4 OR #5 1504

#7 MeSH descriptor: [Vulnerable Populations] this term only 277

#8 MeSH descriptor: [Poverty] explode all trees 1633

#9 (low NEAR/3 income*):ti,ab,kw OR (poor OR poverty OR vulnerabl*):ti,ab,kw OR ((marginal OR precarious* OR disadvantag* OR "at risk") NEAR/3 (people OR youth OR person? OR population? OR individual? OR m?n OR wom?n OR adult?)):ti,ab,kw (Word variations have been searched) 77513

#10 #7 OR #8 OR #9 77513

#11 #6 AND #10 415

#12 #3 OR #11 1564

#13 MeSH descriptor: [Trauma and Stressor Related Disorders] this term only 6

#14 MeSH descriptor: [Stress Disorders, Traumatic] explode all trees 2589

#15 (post-traumatic OR posttraumatic OR ptsd OR traumatic disorder*):ti,ab,kw OR ((posttrauma* OR post-trauma*) NEAR/3 (stress* OR disorder* OR psych* OR symptom*)):ti,ab,kw OR (acute stress disorder* OR combat disorder* OR war neuros*):ti,ab,kw OR (trauma-informed OR trauma-focus*):ti,ab,kw OR (trauma* NEAR/3 (care OR treatment* OR therap* OR intervention*)):ti,ab,kw 10583

#16 #13 OR #14 OR #15 10604

#17 #12 AND #16 61

**PTSDpubs,** formerly known as PILOTS, is a freely available, bibliographic database providing access to the worldwide literature on PTSD and other mental health consequences of traumatic events.

PTSDpubs has unique features that set it apart from other databases. This database offers:

- A custom PTSD and trauma focused thesaurus to help you create a precise search. This unique thesaurus includes specific PTSD symptoms, like hypervigilance, as well as terms such as PTSD (DSM-5) and PTSD (ICD-11) to help you search by diagnostic criteria.
- A detailed listing of tests and measures. Each record in PTSDpubs lists all instruments used within the publication, and you can search for publications that use a specific test or measure.
- A comprehensive range of publication types, including journal articles, books, reports, newsletters, and dissertations.
- Cross-disciplinary coverage of all publications relevant to PTSD and psychological trauma. PTSDpubs does not limit its coverage to selected journals, but tries to include all relevant publications.

~~Search Strategy~~

~~Set#: S11~~

~~Searched for: (MAINSUBJECT.EXACT("Homeless Persons") OR MAINSUBJECT.EXACT("Homelessness") OR MAINSUBJECT.EXACT("Shelter Residents")) OR (homeless* OR "housing outcome" OR "housing outcomes" OR "no home" OR "no homes" OR "no housing" OR evict*) OR (squatter* OR vagrant* OR street-involved OR street person OR street people OR street youth OR "no fixed address" OR "temporary housing" OR "unstably housed" OR "housing instability")~~

~~Databases: PTSDpubs~~

~~Results: 657~~

~~Set#: S13~~

~~Searched for: (MAINSUBJECT.EXACT("Trauma Focused Cognitive Behavioral Therapy") OR MAINSUBJECT.EXACT("Trauma Focused Group Psychotherapy")) OR (trauma-informed OR trauma-focus*) OR (trauma* NEAR/3 (care OR treatment* OR therap* OR intervention*))~~

~~Databases: PTSDpubs~~

~~Results: 5368~~

~~Set#: S14~~

~~Searched for: S11 AND S13~~

~~Databases: PTSDpubs~~

~~These databases are searched for part of your query.~~

~~Results: 41~~

Search Strategy

Set#: S1

Searched for: MAINSUBJECT.EXACT("Homeless Persons") OR MAINSUBJECT.EXACT("Homelessness") OR MAINSUBJECT.EXACT("Shelter Residents")

Databases: PTSDpubs

Results: 499

Set#: S12

Searched for: vulnerab* AND hous*

Databases: PTSDpubs

Results: 50

Set#: S13

Searched for: homeless* OR "no housing" OR "no fixed address" OR squatter* OR evict* OR "housing outcome" OR "housing outcomes" OR "no home" OR "no homes"

Databases: PTSDpubs

Results: 466

Set#: S16

Searched for: (hous* OR home OR homes OR shelter* OR hostel? OR accomodation? OR dwelling?) NEAR/3 (insecur* OR instabil* OR stabil* OR stable OR unstabl* OR temporar* OR marginal* OR precarious* OR inadequate*)

Databases: PTSDpubs

Results: 76

Set#: S17

Searched for: street* NEAR/3 (people OR youth OR person? OR population? OR individual? OR m?n OR wom?n OR adult?)

Databases: PTSDpubs

Results: 31

Set#: S19

Searched for: S1 OR S12 OR S13 OR S16 OR S17

Databases: PTSDpubs

These databases are searched for part of your query.

Results: 719

----------------------------------------------------------
